# A Survey on the Willingness and Demand for Acupuncture Treatment Among Patients with Malignant Tumors

**DOI:** 10.64898/2026.03.24.26349235

**Authors:** Qun Liu, Yun Wang, Yingtian Wang, Sha Luo, Bo Meng, Ye Feng, Zilin Long, Zhandong Li, Dong Xue, Hong Sun

## Abstract

**Objective:** A questionnaire survey was conducted on the willingness and demand for acupuncture treatment in patients with malignant tumors, and the possible factors affecting patients’ willingness and demand for acupuncture treatment were explored.

**Methods:** A voluntary, anonymous survey was conducted between February and May 2025 among patients with malignant tumors aged 18 years and older who visited Beijing Cancer Hospital. The questionnaire included 16 questions addressing three dimensions:current medical purposes,Traditional Chinese Medicine(TCM) literacy, and acupuncture treatment needs.The questionnaire was posted online and completed by respondents using a smartphone interface.

**Results:** A total of 511 valid questionnaires were retrieved in the survey, and 481 patients(94.1%) are willing to receive acupuncture treatment. Among the 481 patients willing to receive acupuncture treatment, the top five symptoms they hoped to improve with acupuncture were: disturbed sleep (245 participants, 50.9%); pain (229 participants, 47.6%); fatigue (177 participants, 36.8%); numbness (165 participants, 34.3%); and poor appetite (144 participants, 29.9%). Among patients who chose to “explicitly accept” acupuncture treatment and those who “accepted acupuncture treatment upon doctor’s recommendation”, 55% and 56% respectively had good knowledge of traditional Chinese medicine (TCM) culture. In contrast, this proportion was only 36.7% among patients who refused acupuncture treatment, and the difference was statistically significant (P<0.05). The survey results also show that Female patients reported significantly higher demands for pain relief and improved sleep than male patients, with statistically significant differences (*P*<0.05). Furthermore, those aged 18-45 and with better TCM literacy were more likely to desire acupuncture to improve sleep, with statistically significant differences (*P*<0.05).

**Conclusion:** Differences in TCM literacy can influence patients’ willingness to choose acupuncture treatment. Strengthening patient health education and improving TCM literacy will help increase cancer patients’ willingness to choose TCM acupuncture treatment, thereby enabling them to benefit from acupuncture. For patients aged 18-45, those with good TCM literacy female with high acupuncture needs, acupuncture treatment may be recommended as a priority.

## 1. Research Introduction

The World Health Organization’s International Agency for Research on Cancer (IARC) released a report stating that in 2022, there were approximately 20 million new cancer cases and about 9.7 million cancer deaths globally^[1] [2]^. In the face of the continuous harm caused by cancer to human health and the social economy, people have made comprehensive efforts to address this challenge, including the prevention of cancer, early screening, comprehensive treatment, psychological support, rehabilitation services, and the exploration and research of new treatment methods.

In traditional Chinese medicine (TCM), it is believed that yang deficiency with cold congelation and qi stagnation with blood stasis are the main pathogenesis of tumor complications. Acupuncture, as a typical TCM therapy, can exert the effects of warming yang and dredging collaterals as well as replenishing qi and activating blood circulation by stimulating acupoints.^[3]^. Numerous studies have shown^[4], [5], [6]^ that Acupuncture treatment can alleviate various uncomfortable symptoms in patients during radiotherapy, chemotherapy, and post-surgical rehabilitation, while improving their quality of life. For instance, its effects in relieving cancer-related pain and cancer-related fatigue, promoting gastrointestinal function in patients after abdominal tumor surgery, and alleviating hand-foot numbness caused by chemotherapy have all been verified in relevant studies^[7][8][9]^and recommended for use in a variety of cancer treatment guidelines.

However, in current real-world scenarios, the application of acupuncture in cancer treatment and rehabilitation remains limited. Studies have found^[10]^ that patients’ lack of understanding of acupuncture, vague perceptions of acupuncture treatment, and lack of identification with acupuncture culture may be factors that influence these patients’ refusal to choose acupuncture treatment. In a survey of acupuncture treatment for common illnesses, such as common pain, bone and joint muscle disorders, and general insomnia and anxiety, the study showed that before the interview, patients indicated they would not consider acupuncture treatment when ill, but after the interview, the number of patients choosing acupuncture treatment increased compared to before the interview. Recommendations from friends and family, trust in traditional Chinese medicine, the hospital’s good brand, and word-of-mouth with doctors are the main reasons patients seek treatment in acupuncture departments. Furthermore, the initial efficacy of acupuncture treatment is consistent with patients’ expectations^[11]^. Therefore, investigating the understanding and needs of malignant tumor patients regarding acupuncture treatment can help provide more accurate treatment recommendations for patients and increase awareness of the therapeutic benefits of acupuncture among cancer patients and medical professionals.

## 2. Methods

### 2.1 Questionnaire Design

Based on the clinical experience of acupuncture therapy for tumor rehabilitation, a self-designed questionnaire was developed by integrating content from the Traditional Chinese Medicine (TCM) version of the MD Anderson Symptom Inventory (MDASI–TCM) and the 2020 Questionnaire on the Health Literacy of Traditional Chinese Medicine among Chinese Citizens. The questionnaire consists of 16 items, covering three dimensions as follows: patients’ purpose of consultation, TCM cultural literacy, and demand for acupuncture therapy.

Based on the data from a pre-survey of 60 questionnaires, the item settings were revised and refined. Using SPSS 29 to test the questionnaire, the Cronbach’s α coefficient for reliability was 0.809, the Kaiser-Meyer-Olkin (KMO) value for sampling adequacy (a measure of validity) was 0.887, and the Bartlett’s Test of Sphericity yielded a p-value < 0.001. This questionnaire demonstrates high stability and reliability, and can accurately reflect the current state of knowledge and usage needs of patients with malignant tumors regarding acupuncture therapy. (Table 1).

**Table 1.**
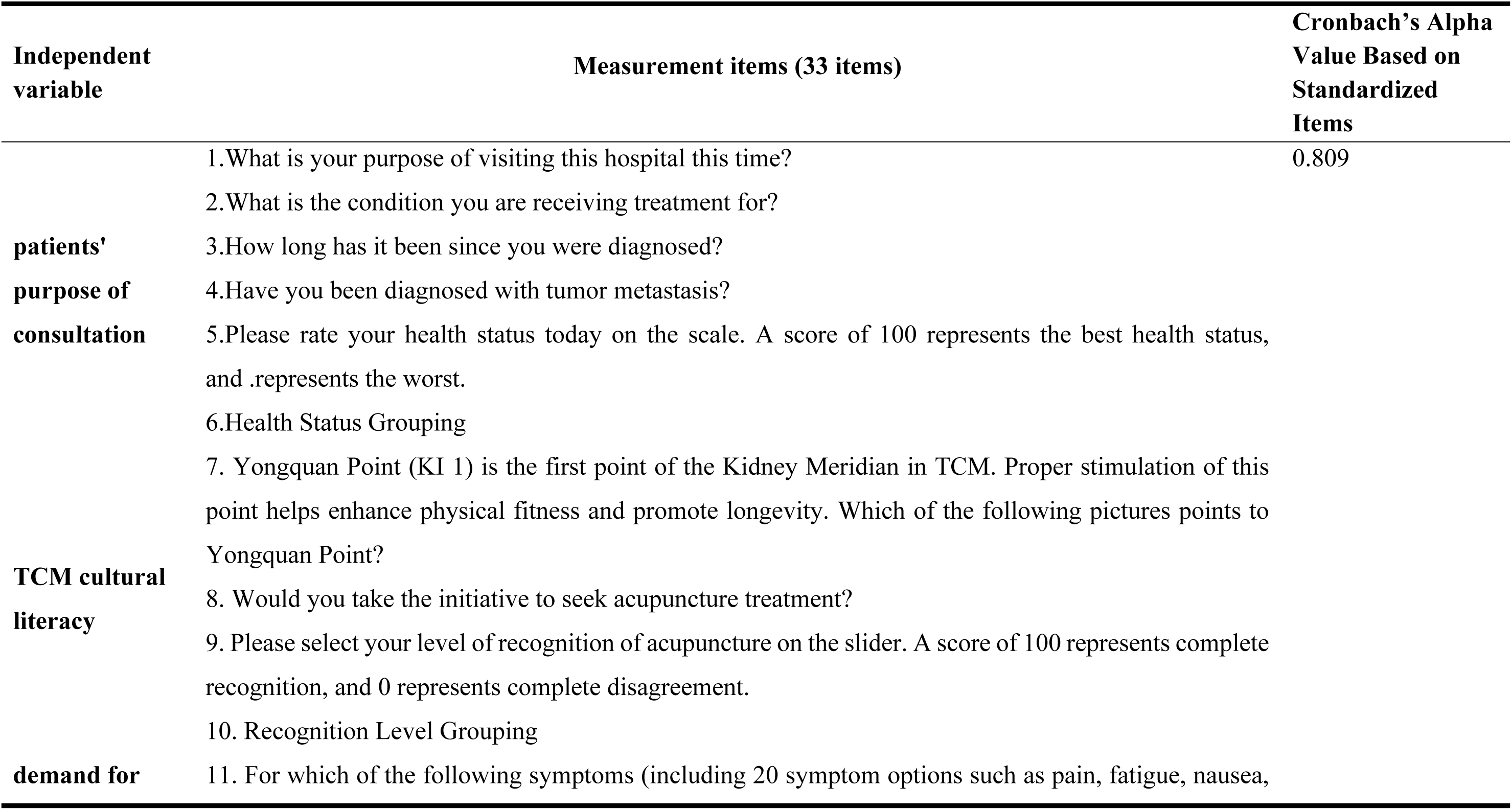
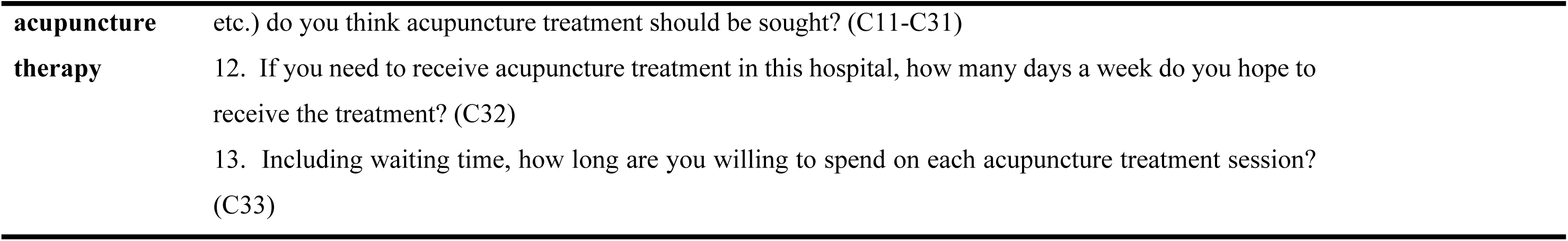
Questionnaire Design and Reliability Analysis.

### 2.2 subjects

The subjects of this study were inpatients and outpatients who visited Peking University Cancer Hospital from February 2025 to April 2025.

Inclusion Criteria:Patients with a pathological diagnosis of malignant tumor;Patients aged over 18 years old.

Exclusion Criteria:Patients with cognitive impairment;Patients with an ECOG(Eastern Cooperative Oncology Group) score greater than 3;Practitioners related to the medical industry.

### 2.3 Survey methods

A convenient sampling method was adopted, and a voluntary, anonymous questionnaire survey was conducted. The questionnaire was created and distributed via Sojump (www.sojump.com), an online survey platform, and respondents completed the questionnaire by filling in information on their smartphone interfaces.

For inpatients, bedside visits were conducted: patients who met the inclusion criteria were informed of the survey’s purpose, significance, methods, and required time. After obtaining their consent, they first signed an informed consent form, then completed the questionnaire. If a patient was unable to complete the questionnaire independently, a family member could fill it out on their behalf, or the patient could dictate the answers while the researcher completed the form. Efforts were made to ensure the completeness of all questionnaires.

For outpatients, questionnaires were collected by posting posters at the information desk and having medical guides direct patients to fill out the questionnaires.

### 2.4 Statistical analysis

SPSS 29 software was used, and the chi-square test was applied to analyze the differences in the demand rate for acupuncture therapy among patient groups with different characteristics. Respondents who held a negative attitude towards acupuncture therapy (i.e., refused it) were guided to participate in patient interviews to elaborate on the specific content of their concerns regarding acupuncture therapy.

### 2.5 Ethical standards

This research protocol has been reviewed and approved by the Ethics Committee of Peking University Cancer Hospital (Approval No.: 2025YJZ06) and complies with ethical requirements.

## 3. Results

### 3.1 General Demographic Characteristics

The survey was conducted from February 24, 2025 to April 30, 2025. A total of 739 questionnaire leaflets were distributed, and 511 valid questionnaires were collected, with an effective response rate of 69.1%. Among the respondents:

- By gender: 247 were male (48.3%) and 264 were female (51.7%).
- By age group: 180 were aged 18-45 years (35.2%), 174 were aged 46-60 years (34.1%), and 162 were over 60 years old (31.7%).
- By type of tumor: 93 had lung cancer (18.2%), 82 had breast cancer (16.1%), 55 had colon cancer (10.8%), 59 had gastric cancer (11.5%), 40 had rectal cancer (7.8%), 21 had esophageal cancer (4.1%), 26 had gynecological tumors (5.1%), 12 had liver cancer (2.3%), and 123 had other types of tumors (24.1%).

In terms of the duration since tumor diagnosis (by the time of the survey):

- 200 respondents (39.1%) were diagnosed 0-3 months prior;
- 95 respondents (18.6%) were diagnosed 4-6 months prior;
- 61 respondents (11.9%) were diagnosed 7-12 months prior;
- 68 respondents (13.3%) were diagnosed 1-2 years prior;
- 50 respondents (9.8%) were diagnosed 3-5 years prior;
- 37 respondents (7.3%) were diagnosed more than 5 years prior.

In terms of tumor stage:

- 228 patients (44.6%) had early-to-mid-stage tumors;
- 210 patients (41.1%) had advanced-stage tumors;
- 73 patients (14.3%) were unsure whether they had tumor metastasis at present. (Table 2).

**Table 2.**
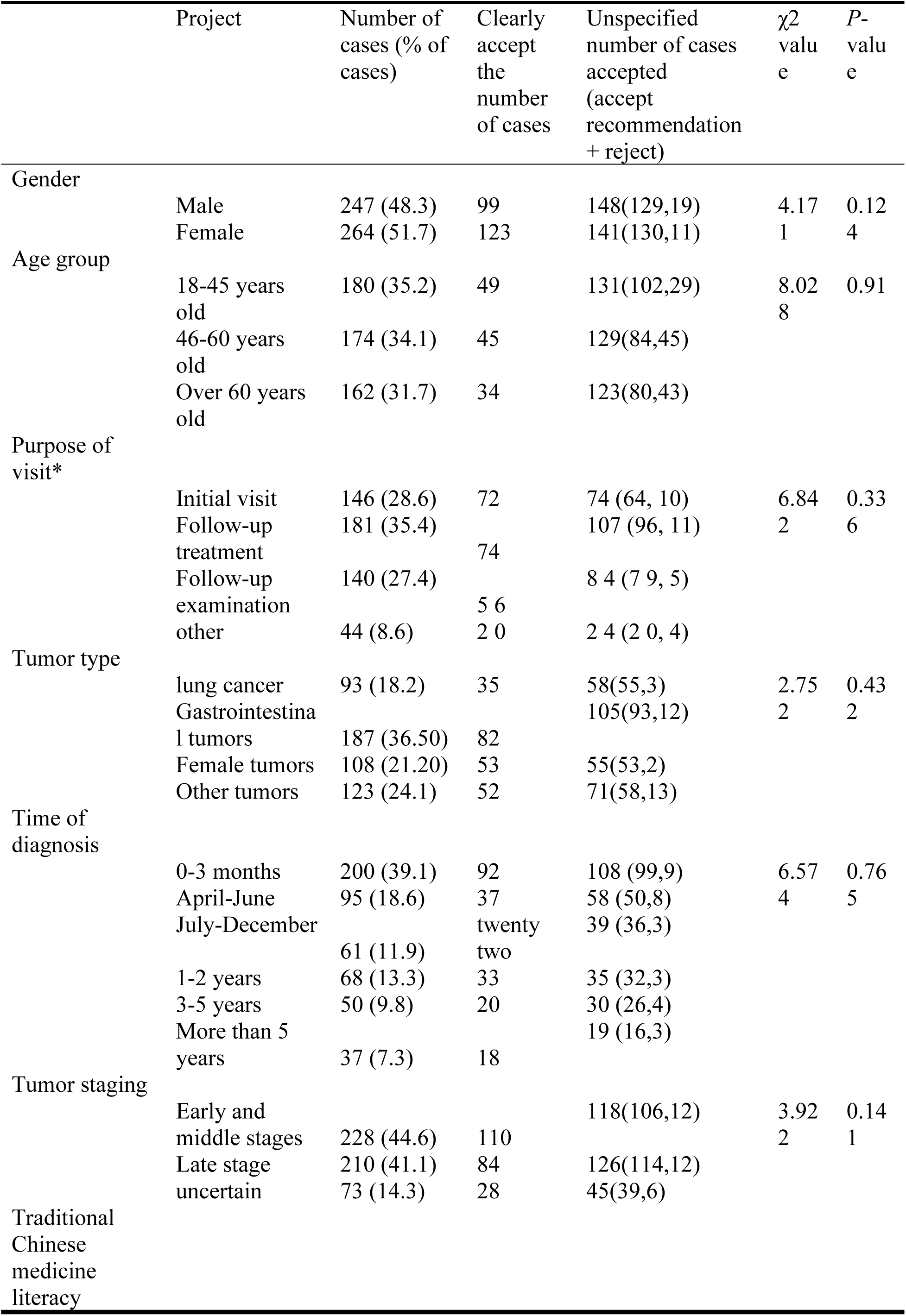
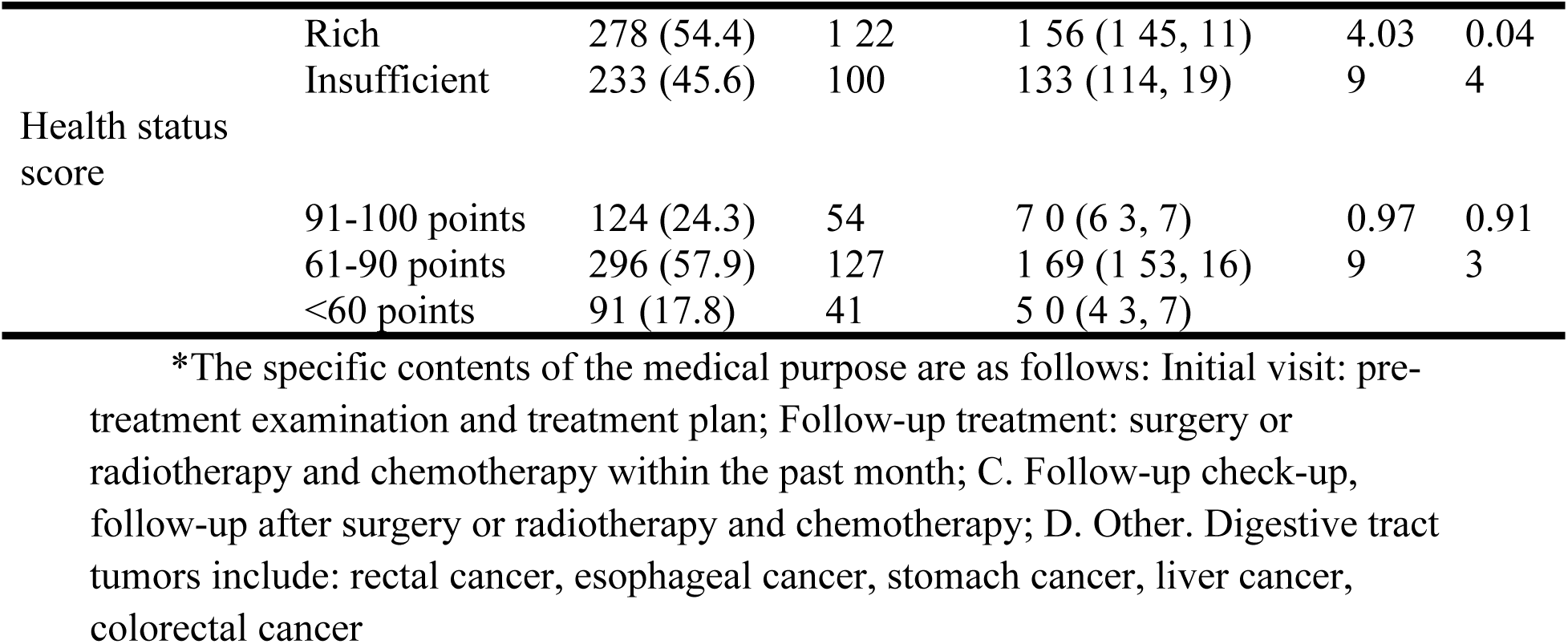
Comparison of General Conditions Among Surveyed Tumor Patients.

### 3.2 Attitudes of Different Patient Groups Toward Choosing TCM Acupuncture Therapy

The survey analyzed the attitudes of different tumor patient groups toward choosing Traditional Chinese Medicine (TCM) acupuncture therapy, focusing on factors such as TCM health literacy, gender, age group, purpose of medical consultation, tumor type, duration since diagnosis, tumor stage, and health status score. Key findings are as follows:

#### 3.2.1 Attitudes Correlated with TCM Health Literacy: More Positive Attitudes in Patients with Higher Literacy

TCM health literacy—defined as patients’ understanding of TCM cultural knowledge and recognition of TCM efficacy—shows a significant correlation with their attitudes toward acupuncture therapy. The specific distribution is as Table 3. Statistical result: χ² = 4.039, P = 0.044.

**Table 3.** Attitudes Correlated with TCM Health Literacy.

| Attitude Toward Acupuncture Therapy | Proportion | Group Characteristics |
| --- | --- | --- |
| Clearly willing to accept | 55% | Patients actively recognize the value of acupuncture and take the initiative to choose it. |
| Acceptable if recommended by doctors | 56% | Patients hold a neutral-to-positive attitude and rely on professional advice to make decisions. |
| Refuse to accept | 36.7% | Patients lack understanding of acupuncture or have doubts about its efficacy. |

The difference in the proportion of patients with good TCM cultural knowledge among the three attitude groups is statistically significant. This indicates that patients with higher TCM health literacy have a more positive perception of acupuncture therapy, while those with lower literacy are more likely to reject it.

#### 3.2.2 Attitudes Correlated with Gender: Higher Willingness in Female Patients

Gender differences in attitudes toward acupuncture therapy were observed, with female patients showing a stronger willingness to accept acupuncture compared to male patients(Table 4).Statistical result: χ² = 3.727, P = 0.054.

**Table 4.** Attitudes Correlated with Gender.

| Attitude Toward Acupuncture Therapy | Female Patients (n=264) | Male Patients (n=247) |
| --- | --- | --- |
| Clearly willing to accept | 46.6% | 40.1% |
| Acceptable if recommended by doctors | 49.3% | 52.2% |
| Refuse to accept | 4.1% | 7.7% |

Although the difference does not reach the level of statistical significance (P > 0.05), the data still reflects a clear trend:

The proportion of female patients who “clearly willing to accept” acupuncture is 6.5 percentage points higher than that of male patients.

The proportion of female patients who “refuse to accept” acupuncture is 3.6 percentage points lower than that of male patients.

This suggests that female patients may have a more open attitude toward acupuncture therapy, which may be related to factors such as women’s higher attention to complementary therapies and better acceptance of non-pharmaceutical interventions.

#### 3.2.3 No Significant Correlation with Other Demographic and Clinical Factors

In addition to TCM health literacy and gender, the survey also analyzed the correlation between attitudes toward acupuncture and other variables. No statistically significant differences were found in the groups. (Table 5)

**Table 5.** Correlation with Other Demographic and Clinical Factors.

| Factor Category | Specific Groupings | Key Finding |
| --- | --- | --- |
| Age group | 18–45 years old, 46–60 years old, >60 years old | The willingness to accept acupuncture does not vary with age; no obvious age-related preference was observed. |
| Purpose of medical consultation | Treatment of tumor-related symptoms (e.g., pain, fatigue), rehabilitation after treatment, symptom relief during radiotherapy/chemotherapy | Attitudes toward acupuncture are not affected by the purpose of consultation; patients with different medical needs show similar acceptance. |
| Tumor type | Lung cancer, breast cancer, gastric cancer, colorectal cancer, gynecological tumors, etc. | There is no significant difference in acupuncture acceptance among patients with different tumor types. |
| Duration since diagnosis | 0–3 months, 4–6 months, 7–12 months, 1–2 years, 3–5 years, >5 years | Newly diagnosed patients and long-term survivors have similar attitudes toward acupuncture; no correlation with disease course was found. |
| Tumor stage | Early-to-mid-stage (I–II), advanced-stage (III–IV) | The acceptance of acupuncture does not differ between early-stage and advanced-stage patients. |
| Health status score | Scored by patients based on subjective physical condition (e.g., 1–10 points) | Patients with different self-rated health statuses show no significant difference in their attitudes toward acupuncture. |

##### Summary of Key Findings

TCM health literacy is a core influencing factor: Patients with good TCM cultural knowledge are significantly more likely to accept acupuncture therapy, while those with low literacy tend to refuse it. This highlights the importance of popularizing TCM knowledge to improve patients’ acceptance of complementary therapies.

Gender shows a potential trend: Although not statistically significant, female patients have a higher willingness to accept acupuncture than male patients, which may provide a reference for targeted communication when promoting acupuncture therapy. No correlation with other clinical/demographic factors: Variables such as age, tumor type, and stage do not affect patients’ attitudes toward acupuncture, indicating that acupuncture therapy has broad acceptance potential across different patient groups.

### 3.3 Characteristics of patients’ acupuncture needs

Among 481 patients willing to receive acupuncture treatment, the top 5 symptoms they hoped to improve through acupuncture (i.e., the top 5 acupuncture treatment needs) were as follows: 245 patients (50.9%) had sleep disturbance; 229 patients (47.6%) had pain; 177 patients (36.8%) had fatigue; 165 patients (34.3%) had numbness; and 144 patients (29.9%) had poor appetite.

In terms of treatment frequency and duration, 84.6% of the patients were willing to receive acupuncture 1 to 3 times per week, and 49.9% were willing to spend 30 minutes per acupuncture session. (Table 6).

**Table 6.** Characteristics of Patients’ Acupuncture Needs.

| Feature | Number of cases | Proportion (%) |
| --- | --- | --- |
| What discomfort symptoms should be treated with acupuncture? |  |  |
| Disturbed sleep | 245 | 50.9 |
| Pain | 229 | 47.6 |
| Fatigue | 177 | 36.8 |
| Numbness | 165 | 34.3 |
| Poor appetite | 144 | 29.9 |
| Nausea | 120 | 24.9 |
| Shortness of breath | 111 | 23.1 |
| Upset | 111 | 23.1 |
| Constipate | 103 | 21.4 |
| Cough | 87 | 18.1 |
| + Hot palms and soles | 83 | 17.3 |
| How many times a week would you like to receive acupuncture treatments? |  |  |
| 1 day | 177 | 36.8 |
| 2 days | 111 | 23.1 |
| 3 days | 119 | 24.7 |
| 4 days | 13 | 2.7 |
| 5 days | 29 | 6 |
| 6 days | 9 | 1.9 |
| 7 days | 23 | 4.8 |
| How long would you like to spend on each acupuncture treatment? |  |  |
| 15 minutes | 80 | 16.6 |
| 30 minutes | 240 | 49.9 |
| 60 minutes | 116 | 24.1 |
| More than 1h | 45 | 9.4 |

In terms of the demand for acupuncture treatment among different populations:

Female patients had significantly higher demands for pain relief and sleep improvement than male patients, with a statistically significant difference (*P* <0.05).

Among patients of different age groups, those aged 18-45 were more likely to hope to improve sleep through acupuncture (*P* <0.05).

Among patients with different levels of TCM literacy, those with better TCM literacy were more inclined to expect sleep improvement via acupuncture (*P* <0.05). (*P*<0.05) (Table 7).

**Table 7.** Analysis of Differences in Acupuncture Treatment Needs Among Different Populations.

| Project |  | Number of cases<br>(percentage) | Pain relief needs |  |  |  | Reduce fatigue needs |  |  |  | Improve sleep needs |  |  |  |
| --- | --- | --- | --- | --- | --- | --- | --- | --- | --- | --- | --- | --- | --- | --- |
| | | | Yes | None | $\chi^2$ value | *P* value | Yes | None | $\chi^2$ value | *P*-value | Yes | None | $\chi^2$ value | *P*-value |
| Gender | Male | 2 28 (4 7.4 %) | 9 7 | 1 31 | 4.459 | 0.035 | 78 | 1 50 | 1.248 | 0.264 | 99 | 129 | 9.794 | 0.002 |
|  | Female | 2 53 (5 2.6 %) | 1 32 | 1 21 |  |  | 99 | 1 54 |  |  | 146 | 107 |  |  |
| Age group | 18-45 years old | 1 72 (3 5.8 %) | 8 5 | 8 7 | 2.931 | 0.231 | 6 4 | 1 08 | 1.776 | 0.412 | 1 02 | 7 0 | 8.160 | 0.017 |
|  | 46-60 years old | 1 63 (3 3.9%) | 8 3 | 8 0 |  |  | 5 4 | 1 09 |  |  | 7 9 | 8 4 |  |  |
|  | Over 60 years old | 1 46 (3 0.3%) | 6 1 | 8 5 |  |  | 5 9 | 8 7 |  |  | 6 4 | 8 2 |  |  |
| TCM literacy | Rich | 2 67 (5 5.5 %) | 1 39 | 1 28 | 4.972 | 0.083 | 9 7 | 1 70 | 0.875 | 0.645 | 1 57 | 1 10 | 1 8.483 | < 0.001 |
|  | Insufficient | 214 (44.5 %) | 90 | 124 |  |  | 8 0 | 1 34 |  |  | 8 8 | 1 26 |  |  |

## 4. Discussion

This study investigates the willingness and demand for acupuncture treatment among patients with malignant tumors, while also exploring the role of Traditional Chinese Medicine cultural literacy. Similar to the findings of previous studies^[12] [13]14^,most participants indicated that they would be willing to try the therapy after being informed of the potential benefft of acupuncture for alleviate the side effects of chemotherapy.

More importantly, the survey data indicate three trendst as follows:firstly,participants with different attitudes toward acupuncture exhibit varying levels of TCM literacy;Secondly,they would be willing to try the therapy after being informed of the potential benefft by Oncologists; lastly,there are differences in acupuncture treatment needs among different cancer patient groups. The three points mentioned above make the topic holds more considerable clinical relevance and practical value. However,due to limitations in personal capacity and time, this survey was conducted solely at Beijing Cancer Hospital, and the findings are inevitably subject to bias. For future surveys, probability sampling methods such as stratified random sampling and systematic sampling, or quota sampling, can be adopted to reduce such bias.

### 4.1 Willingness of Cancer Patients to Receive Acupuncture Treatment Is Influenced by Their Literacy in Traditional Chinese Medicine (TCM) Culture

In recent years, modern medicine has achieved numerous outcomes in researching the side effects of anti-tumor treatments. Meanwhile, acupuncture has also gained a wealth of effective methods in this regard, and its role in cancer treatment has attracted increasing attention. In China, Traditional Chinese Medicine (TCM) acupuncture is a therapeutic approach that plays an important role in anti-tumor treatment. Patients with a high level of TCM cultural literacy, while receiving modern medical anti-tumor treatment, actively choose TCM acupuncture to enhance their own immunity, reduce treatment side effects, thereby achieving a better quality of life and avoiding suffering from symptoms such as pain, fatigue, nausea, and vomiting.

In this questionnaire study, patients’ TCM cultural literacy was evaluated through questions on meridian and acupoint identification. The analysis results showed that patients with high TCM cultural literacy held a more positive attitude towards choosing TCM acupuncture treatment. Patients with higher TCM cultural literacy often have access to more information about acupuncture treatment and attach equal importance to traditional health preservation and modern diagnosis and treatment. Therefore, the proportion of patients who either explicitly accept acupuncture treatment or are willing to follow doctors’ recommendations is much higher than that of patients with low TCM cultural literacy.

Among the respondents in this study, 278 patients (accounting for 54.4%) had relatively high TCM literacy, but the general population still lacks the promotion of TCM knowledge. Therefore, in the process of cancer prevention and treatment, improving the public’s TCM literacy can increase cancer patients’ willingness to choose TCM acupuncture treatment and enhance their quality of life.

### 4.2 Recommendations from Oncologists Can Increase Patients’ Willingness to Receive Acupuncture Treatment

For patients with malignant tumors, recommendations from oncologists can help alleviate patients’ concerns about safety and play a crucial role in promoting patients’ choice of acupuncture treatment. When patients learn from their specialists that acupuncture can be used as an adjuvant therapy to benefit disease recovery, they are more likely to accept acupuncture treatment. Currently, research institutions such as the Affiliated Hospital of Beijing University of Chinese Medicine (China), the Affiliated Hospital of Guangzhou University of Chinese Medicine (China), and Memorial Sloan Kettering Cancer Center (USA) have been conducting large-scale, continuous research on acupuncture for cancer treatment. Cancer patients in these hospitals have received high-quality acupuncture treatment.

From the perspective of evidence-based medicine, presenting the research results of acupuncture in the field of cancer treatment at major international oncology conferences helps enhance the influence of acupuncture treatment^[15][16][ 17]^.It also provides strong evidence support for oncologists and developers of cancer treatment guidelines. For example, the National Comprehensive Cancer Network (NCCN) recommends acupuncture or electroacupuncture as a non-pharmacological intervention for cancer pain management in its Adult Cancer Pain Guidelines (Version 2025.1). Another example is the Annual Meeting of the American Society of Clinical Oncology (ASCO), which is regarded as one of the world’s most important oncology conferences. The latest research findings and clinical data released at this conference play a significant guiding role in the development of cancer treatment. At the 2024 Annual Meeting, a study on acupuncture for chemotherapy-induced peripheral neuropathy (CIPN) was published^[18][19]^. The results showed that an 8-week acupuncture protocol significantly improved the CIPN-related quality of life in women with early-stage breast cancer induced by paclitaxel, which has important clinical significance. In China, the Progress in Cancer Acupuncture Research released by the China Anti-Cancer Association (CACA) summarizes the progress of clinical and preclinical research on cancer acupuncture in China over the past three years, including clinical research in 8 areas and a number of preclinical studies. This has played a positive role in promoting the widespread application of cancer acupuncture treatment in China.

### 4.3 Differences in Acupuncture Treatment Needs Among Different Cancer Patient Groups

The survey found that female patients aged 18–45 with good TCM literacy had the highest demand for sleep improvement. Meanwhile, female patients also had a significantly higher demand for pain relief than male patients. Some acupuncture experts have pointed out that patients’ understanding of acupuncture has a significant impact on the therapeutic effect of acupuncture^[20]^. Middle-aged and young patients have a strong need to actively seek relief from psychological symptoms such as anxiety, insomnia, and depression, and this factor can help this group achieve better therapeutic effects through acupuncture treatment.

In fact, there is abundant research evidence on the role of acupuncture in improving sleep and relieving pain. Studies on acupuncture for sleep improvement^[21][22]^have shown that acupuncture can improve sleep quality in cancer patients with insomnia without causing serious adverse events, especially among Asian participants. A subgroup meta-analysis revealed that after 6 to 8 weeks of acupuncture treatment, acupuncture was superior to the wait-list control group in improving the total score of the Pittsburgh Sleep Quality Index (PSQI). Among breast cancer patients undergoing cancer treatment, acupuncture achieved a higher effective rate than benzodiazepines at the 1-week follow-up after treatment. Research on acupuncture for pain relief^[23]^ has demonstrated that the integration of acupuncture with analgesic drugs can significantly alleviate pain and ameliorate adverse effects of analgesics in patients with chronic cancer pain, thereby improving patients’ quality of life^[24] [25]26^. Therefore, for female patients troubled by pain and sleep disorders, acupuncture treatment can be prioritized as a recommended option.

In addition, patients undergoing radiotherapy, chemotherapy, or surgery all tend to choose acupuncture to treat nausea and loss of appetite. The occurrence of gastrointestinal symptoms during such anti-tumor treatments, if persistent for a long time, can lead to metabolic disorders, malnutrition, dehydration, and immune deficiency. These are important factors that cause anti-cancer treatments to fail to achieve expected results and even accelerate disease progression. Numerous studies have confirmed that acupuncture has a clear therapeutic effect on cancer-related nausea and vomiting, as it can significantly reduce the severity of symptoms, shorten the duration of symptoms, and improve patients’ quality of life.

### 4.4 Limitations of This Survey Study

All participants in this survey were patients attending Beijing Cancer Hospital, and simple random sampling was adopted. Therefore, this survey has considerable limitations, as it only reflects the characteristics of a subset of patients with higher educational levels, better socioeconomic status, and greater exposure to Traditional Chinese Medicine (TCM).

The results of this survey are only intended to improve TCM acupuncture-related services for patients at Beijing Cancer Hospital, and they neither represent nor are applicable to patients in primary medical institutions in other cities or rural areas, nor to community-based healthy populations.Therefore, future studies involving larger and more diverse populations are essential.

## 5. Conclusions

Cancer patients’ willingness to receive acupuncture treatment is significantly influenced by their TCM cultural literacy. Patients with a higher level of TCM cultural literacy hold a more positive attitude toward choosing TCM acupuncture treatment, and their acceptance of acupuncture is much higher than that of patients with low TCM cultural literacy.

Different patient groups have different needs for acupuncture treatment. Among them, female patients aged 18–45 with good TCM literacy have the highest demand for sleep improvement. Meanwhile, female patients also have a significantly higher demand for pain relief than male patients. Therefore, for female patients troubled by pain and sleep disorders, acupuncture treatment can be prioritized.

Currently, the general cancer patient population lacks knowledge of TCM, and the level of TCM health literacy remains low. In the process of cancer prevention and treatment, strengthening health education for patients and improving the population’s TCM literacy will help increase cancer patients’ willingness to choose TCM acupuncture treatment, thereby enabling them to benefit from acupuncture therapy.

## Acknowledgments

We would like to thank all the individuals and institutions that have contrib uted to this work.

## Authors’ contributions

Qun Liu and Yun Wang contributed equally to this work. Qun Liu and Yun Wang designed the study, collected data, drafted the manuscript and tables.Yingtian,Sha Luo,Bo Meng,Ye Feng and Zilin Long participated in data collection, manuscript revision. Zhandong Li,Dong Xue and Hong Sun conceived and supervised the study, provided critical intellectual input, and approved the final version of the manuscript. All authors read and approved the final manuscript.

## Data availability

All data generated during this study are included in this manuscript.

## Declarations

### Ethics approval and consent to participate

Not applicable.

## Consent for publication

Individual patient consent was not applicable as no information in this article are identifiable.

## Funding

Beijing Cancer Hospital Clinical Research Youth Fund. Project Number LGH2024489

Integrative Chinese and Western Medicine Bladder Cancer Professional Committee of China Anti-Cancer Association

